# Biomarker Signal Architecture in Cardiovascular Machine Learning: Stability, Redundancy, and Minimal High-Yield Panels After Myocardial Infarction

**DOI:** 10.64898/2026.05.19.26353638

**Authors:** Natalia Piórkowska¹, Agnieszka Olejnik, Alan Ostromęcki, Wiktor Kuliczkowski, Andrzej Mysiak, Iwona Bil-Lula

**Affiliations:** Faculty of Information and Communication Technology, Wroclaw University of Science and Technology, Wrocław, Poland; Department of Medical Laboratory Diagnostics, Division of Clinical Chemistry and Laboratory Haematology, Faculty of Pharmacy, Wroclaw Medical University, Wroclaw, Poland; Independent Researcher; Institute of Heart Diseases, Wroclaw Medical University, Wroclaw, Poland; Specialized Immunology Laboratory, Clinical Department of Nephrology, Transplantation Medicine and Internal Diseases, Institute of Internal Diseases, Wroclaw Medical University, Poland

**Keywords:** machine learning, myocardial infarction, biomarkers, feature importance, redundancy, complementarity, signal architecture, cardiovascular prediction

## Abstract

**Background:** Machine-learning models based on circulating biomarkers are increasingly used in cardiovascular research; however, model performance alone provides limited insight into how the predictive signal is distributed across features. We aimed to characterize the biomarker signal architecture of a machine-learning model distinguishing ST-elevation myocardial infarction (STEMI) from non-ST-elevation myocardial infarction (NSTEMI), with a focus on signal concentration, redundancy, and conditional complementarity.

**Methods:** We conducted a structured secondary analysis of a previously established, leakage-controlled machine-learning framework (n = 152 patients). The BIOMARKERS feature-set variant (10 biomarkers) was evaluated using outer-fold cross-validation. Model structure was interrogated using (i) leave-one-biomarker-out analysis, (ii) pairwise leave-two-out analysis with pair-excess estimation, (iii) cumulative ablation of top-ranked biomarkers, and (iv) forward reconstruction of minimal biomarker panels. Uncertainty was assessed using bootstrap resampling across folds.

**Results:** The full biomarker model achieved a mean ROC-AUC approaching 0.94. The predictive signal was highly non-uniform, with MMP-2 showing the largest single-feature contribution (mean ΔAUC ≈ 0.16). Pairwise analysis identified conditional complementarity between selected non-lipid biomarkers, particularly MMP-2 and EMMPRIN (pair ΔAUC ≈ 0.26; positive excess over single-feature effects), whereas lipid-related markers formed a highly correlated and largely redundant sub-cluster. Cumulative ablation demonstrated rapid performance collapse following removal of top-ranked biomarkers, consistent with structural signal concentration. Forward panel analysis showed that a compact subset of biomarkers (three features) achieved performance within ∼0.01 ROC-AUC of the full model, indicating the presence of a minimal high-yield panel. Bootstrap confidence intervals suggested that small performance differences should be interpreted with caution.

**Conclusions:** Predictive performance in this biomarker-based model arises from a structured and unevenly distributed signal architecture, characterized by a dominant core biomarker, conditionally complementary contributors, and a redundant lipid cluster. These findings highlight the importance of evaluating model structure, not only aggregate performance, and suggest that biomarker-based machine-learning systems may benefit from architecture-aware interpretation and simplification strategies.

## Introduction

One of the major clinical manifestations of cardiovascular disease is acute myocardial infarction (MI); however, it is not a biologically uniform condition. ST-elevation myocardial infarction (STEMI) and non-ST-elevation myocardial infarction (NSTEMI) are both caused by the coronary artery occlusion. They are usually separated clinically by their electrocardiographic presentation and therapeutic pathways [1]. The conventional STEMI/NSTEMI classification is clinically useful but does not fully explain the differences in inflammation, extracellular matrix remodelling, metabolic stress, lipid metabolism, or endothelial activation. Both MI types arise from complex, partially overlapping biological processes, in which circulating biomarkers may reflect different phases of myocardial injury and systemic response [2,3]. Recent literature on biomarkers in MI indicates that, in addition to troponins, there is growing interest in markers of inflammation, vascular damage, remodelling, and prognostic. Still, their role in multivariate models remains difficult to interpret [1,4,5]. Circulating biomarkers are helpful for cardiovascular prediction because they are relatively accessible, quantitative, and biologically interpretable. However, single biomarkers rarely reflect the full complexity of the tissue’s response after MI [6]. Many biomarkers are biologically or statistically related to each other, e.g. lipid, inflammatory or remodelling markers [6,7]. The main question in biomarker-based models is therefore not only whether a marker is associated with the outcome, but whether it contributes information that is independent, redundant, or conditionally useful in the presence of other factors.

Machine-learning (ML) methods are increasingly used in cardiovascular research because they can model nonlinear associations and interactions among clinical and laboratory variables [8,9]. In recent years, several studies have applied ML for diagnosis, risk prediction and identification of biomarkers in cardiovascular diseases [9–11]. To identify potentially relevant predictors, interpretable ML and feature-importance methods are now commonly used, but they often limit model interpretation to a ranking of variables [12]. High predictive accuracy is not equivalent to mechanistic transparency or structural understanding of how the model uses information. ML analyses often identify features with the greatest predictive value; they are less likely to provide answers to why a given variable is important and whether its role stems from independent contributions, redundancy, or complementarity with other features [7,13]. This is particularly important in biomarker analysis, as correlated variables may capture overlapping biological information, and the contribution of some markers may only become apparent within a specific multivariate context [7,11]. Therefore, the interpretation of ML models using biomarkers should go beyond the simple identification of the most relevant predictors and focus on characterising the internal organisation of the predictive signal, including its concentration, redundancy, and complementarity between individual biomarkers [7,12,13].

The biological heterogeneity of MI is reflected in the involvement of multiple, partially overlapping molecular pathways, including extracellular matrix remodelling, inflammation, lipid metabolism, and mineral–vascular regulation [14–16]. In MI, extracellular matrix remodelling is a central component of the tissue response, with matrix metalloproteinase-2 (MMP-2) [16–18] and extracellular matrix metalloproteinase inducer (EMMPRIN) [19] playing a critical role in post-infarction adaptation and myocardial dysfunction. The level of MMP-2 increases significantly after MI, contributing to collagen and structural protein degradation, inflammatory responses, cardiac rupture, remodelling, and heart failure [16,18]. Inflammatory markers such as interleukin-6 (IL-6) and tumour necrosis factor-alpha (TNF-α) may reflect systemic immune activation. Elevated post-MI IL-6 [20–22] and TNF-α levels [22,23] were related to recurrent coronary events, adverse cardiac remodelling, and increased mortality. Lipids also play a critical, causative role in MI, with low-density lipoprotein cholesterol (LDL) and triglycerides significantly increasing risk [24,25]. As a result of MI, lipid levels often shift, leading to lowered high-density lipoprotein cholesterol (HDL) and altered total cholesterol (TC) levels [26]. Lipid-related biomarkers remain clinically relevant but may also form redundant substructures because several lipid measures encode overlapping metabolic information, especially TC, LDL and non-HDL cholesterol [27]. Klotho and fibroblast growth factor (FGF23) are proteins related to the mineral-vascular axis and biological ageing. They have been recognised as emerging candidate biomarkers in the diagnosis of MI [28–30]. Thus, these biomarkers represent biologically plausible but potentially interdependent sources of predictive information, making them suitable for investigating not only individual biomarker importance but also redundancy, complementarity, and signal organisation within a multivariable ML framework.

Biomarker panels are often proposed to improve diagnostic or prognostic accuracy, but they may contain statistically redundant variables [6,7]. The apparent complexity of a model may therefore be inflated by the redundancy, without improving discrimination [7]. However, eliminating all variables that exhibit similarity or partial correlation may be an oversimplification, because some features, despite sharing biological or statistical information, may still make unique, complementary contributions to a multivariate model [6,7]. Therefore, a clinically meaningful biomarker panel should be evaluated not only in terms of overall predictive performance, but also by its internal signal organisation. In many studies, the discrimination metrics and identification of top-ranked predictors are reported, but formal perturbation-based interpretability analyses, including remove-and-retrain/permutation-based assessments, remains uncommon in cardiology [12,13]. There are works using feature selection and ML for biomarker discovery in MI; however, their primary aim has typically been biomarker identification or optimization of predictive accuracy, rather than to describe the structure of relationships between biomarkers [11,31,32]. Interpretable ML studies frequently use SHAP, permutation importance or related tools, but feature-importance rankings may not fully distinguish dominant, redundant and conditionally complementary predictors [7,12,33,34]. In this context, the present analysis was designed to interrogate the structure of predictive information within a previously established biomarker-based ML framework [35], instead of proposing a new clinical classifier.

This study aimed to characterise the biomarker signal architecture by assessing its concentration, redundancy between variables, conditional complementarity of selected biomarkers and the possibility of isolating a minimal panel of biomarkers retaining high predictive value. The analysis focused on a predefined biomarker-based ML model for STEMI versus NSTEMI classification. The feature set includes markers of extracellular matrix remodelling, inflammation, lipid metabolism, and ageing-related pathways. This framework is intended to improve the interpretation of biomarker-based ML models and to support more transparent development of future cardiovascular biomarker panels.

## 2. Methods

### 2.1 Study design and analytical objective

This study was designed as a methodological secondary analysis of a previously established post-MI ML framework, with the specific aim of characterizing the biomarker signal architecture within the BIOMARKERS feature-set variant. Rather than developing a new prediction model for clinical deployment, the present analysis aimed to determine how the predictive signal was structurally distributed across biomarkers, including the extent of signal concentration, redundancy, conditional complementarity, and the existence of a minimal high-yield biomarker panel.

The analytical strategy was therefore explicitly diagnostic and explanatory at the model-structure level. The focus was not limited to identifying which biomarkers were associated with greater model reliance, but extended to examining whether predictive contribution was concentrated in a small subset of variables, whether some biomarkers behaved as redundant cluster members, and whether specific biomarker pairs contributed complementary information beyond their individual effects.

### 2.2 Data source and study population

The analysis was performed using the same anonymized clinical dataset as in the primary proof-of-concept workflow. The dataset comprised 152 patients with myocardial infarction and included laboratory and clinical variables collected during hospitalization. For the present study, only the predefined BIOMARKERS variant was used. This variant contained 10 biomarker variables representing inflammatory, extracellular matrix remodeling, lipid-related, and aging-related pathways.

The predictive task involved binary classification of STEMI versus NSTEMI, with STEMI treated as the positive class. Only records belonging to labels prespecified as valid for the original task definition were retained. No additional patient selection, subgroup restriction, or post hoc redefinition of the analytical cohort was performed for the present biomarker-architecture analyses.

### 2.3 Predefined biomarker feature set

All analyses in this study were restricted to the biomarker-only feature set predefined during the earlier governance stage of the project. No new feature engineering, feature filtering, or data-driven reclassification of predictors was introduced. The analyzed biomarker panel consisted of the following biomarkers: MMP-2, TC, EMMPRIN, HDL, IL-6, LDL, non-HDL cholesterol, TNF-α, FGF-23, and Klotho.

Feature names were harmonized for reporting purposes only. The biomarker feature set was treated as frozen prior to the analyses reported here.

### 2.4 Analytical framework and leakage control

All analyses were conducted within the same leakage-aware outer-fold structure used in the original nested cross-validation benchmarking workflow. Specifically, the outer stratified folds defined previously for the BIOMARKERS variant were reused without modification. Model fitting, preprocessing, and performance estimation were always conducted separately within each outer-fold split, thereby preserving strict separation between training and held-out test data.

For each outer fold, the best-performing model architecture previously identified for the BIOMARKERS variant was reconstructed using the fold-specific best hyperparameters obtained during the original nested cross-validation procedure. Accordingly, the present analyses did not involve re-running model selection or tuning feature subsets against the full dataset. Instead, they evaluated structured perturbations of the biomarker set under the same fold-specific model specification used in the original workflow.

All preprocessing steps were implemented within scikit-learn pipelines and fitted exclusively on training data on training data within each fold. Median imputation was used for all models, and scaling was additionally applied when required by linear models. This design ensured that feature perturbation analyses remained nested within the original leakage-aware validation framework.

### 2.5 Model reconstruction

For each outer fold, the fold-specific best-performing model for the BIOMARKERS variant was rebuilt using the algorithm selected in the original benchmarking stage and the corresponding fold-level hyperparameters. Depending on the selected best-performing model, the classifier was instantiated as one of the following: Random Forest, Logistic Regression, linear-kernel Support Vector Machine with probability estimation enabled, or HistGradientBoostingClassifier. Random Forest models were reconstructed with a fixed number of 500 trees to match the project’s predefined model specification.

The purpose of model reconstruction in the present study was not to compare model classes, but to preserve consistency with the original validation framework while performing structured biomarker perturbation analyses.

### 2.6 Correlation-based redundancy analysis

Pairwise monotonic associations between biomarkers were quantified using Spearman rank correlation coefficients computed on the full biomarker matrix. To facilitate visualization of the correlation structure, a correlation heatmap was generated following hierarchical clustering based on the distance metric 1 − |ρ|, where ρ denotes the Spearman correlation coefficient. This clustering was used exclusively to reorder variables for visualization and did not alter any downstream modeling analyses.

For interpretative purposes, pairwise redundancy strength was categorized according to the absolute Spearman correlation coefficient as follows: high redundancy for |ρ| ≥ 0.80, moderate redundancy for |ρ| ≥ 0.50, and low redundancy otherwise. These thresholds were used as descriptive aids and were not treated as inferential cutoffs.

### 2.7 Leave-one-biomarker-out analysis

To quantify model dependence on individual biomarkers, a leave-one-biomarker-out analysis was conducted. For each biomarker, the model was re-estimated within each outer fold after removing the corresponding biomarker from the feature set while keeping all remaining biomarkers unchanged. Fold-level ROC-AUC values were then compared with the corresponding outer-fold baseline ROC-AUC obtained from the full BIOMARKERS model.

For each biomarker, the primary summary statistic was the mean fold-level performance drop:

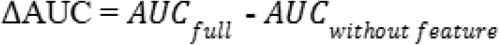

Positive values indicated that removal of the biomarker reduced discrimination and were interpreted as evidence that the biomarker contributed useful predictive information within the model context. Negative values indicated that biomarker removal improved performance on average and were interpreted cautiously as consistent with a potentially noisy or non-contributory role.

Additional descriptive summaries included the median, standard deviation, interquartile range, proportion of folds with any positive performance loss, and proportion of folds with a loss greater than 0.01 ROC-AUC units.

### 2.8 Pairwise leave-two-out analysis

To assess whether pairs of biomarkers behaved as redundant or conditionally complementary predictors, a leave-two-biomarkers-out analysis was performed for all unique biomarker pairs. For each pair, both biomarkers were simultaneously removed, the model was re-estimated within each outer fold, and fold-level ROC-AUC reductions relative to the full BIOMARKERS model were computed.

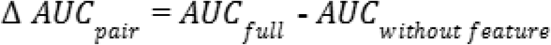

The mean pairwise performance loss was summarized as:

To quantify whether simultaneous removal of a pair caused more harm than expected from the stronger of the two individual effects, the following statistic was calculated:

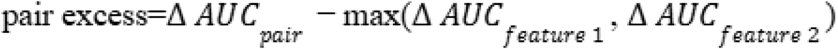

Positive pair-excess values were interpreted as evidence of conditional complementarity, indicating that the pair may contribute information beyond the stronger individual feature effect. In contrast, high pairwise correlation combined with near-zero pair excess was interpreted as consistent with redundancy.

For descriptive interpretation, biomarker pairs were categorized as:

● likely redundant pair, when absolute correlation was high and pair excess was negligible;
● potential complementary pair, when pair excess was clearly positive;
● partially overlapping pair, otherwise.

These labels were used as operational descriptors rather than formal causal claims.

### 2.9 Cumulative ablation analysis

To examine structural signal concentration, a cumulative ablation analysis was performed. Biomarkers were ordered according to the precomputed S12 stability ranking, which was treated as a fixed diagnostic ranking established before the present S14 analyses. This ranking was not recalculated during cumulative ablation and was not optimized against S14 outcomes.

Starting from the highest-ranked biomarker, the top kkk biomarkers were sequentially removed, where k=1,2,…,p −1 and p denotes the total number of biomarkers. For each ablation level, the model was re-estimated within each outer fold using the remaining biomarkers. Mean ROC-AUC, PR-AUC, and Brier scores were then summarized across folds.

The primary quantities of interest were:

● mean ROC-AUC under cumulative removal,
● relative to the full BIOMARKERS model,
● corresponding changes in PR-AUC and Brier score.

This analysis was intended as a structural stress test of how rapidly model performance deteriorated as top-ranked biomarkers were removed, thereby providing indirect evidence about signal concentration in a compact subset of predictors.

For descriptive interpretation, the severity of model collapse was categorized according to the mean ROC-AUC loss versus the full model: minor (<0.03), moderate (0.03 to <0.10), major (0.10 to <0.20), and severe (≥0.20). These categories were used only to summarize the magnitude of degradation.

### 2.10 Forward minimal-panel analysis

To assess whether most predictive signals could be recovered with a compact biomarker subset, a forward panel reconstruction analysis was conducted using the same fixed S12 ranking. Starting from the top-ranked biomarker alone, increasingly larger panels containing the top k biomarkers were evaluated for k=1,2,…,p.

For each panel size, the model was trained and evaluated within the original outer folds, and fold-level ROC-AUC, PR-AUC, and Brier scores were recorded. The principal summary statistics included:

● mean ROC-AUC for the top-k panel,
● ROC-AUC gap relative to the full BIOMARKERS model,
● corresponding PR-AUC and Brier-score gaps.

Panels whose mean ROC-AUC remained within 0.01 or 0.02 of the full BIOMARKERS model were descriptively identified as minimal high-yield panels. These thresholds were prespecified as pragmatic proximity criteria and were not intended to define a clinically optimal panel.

### 2.11 Operational definition of biomarker signal roles

To reduce post hoc interpretative ambiguity, biomarkers were assigned descriptive signal-role categories based on a combination of individual performance loss, correlation structure, pairwise complementarity, and stability information. These categories were intended as operational summaries of model behavior rather than representations of biological ground truth.

The following descriptive roles were used:

● core driver: biomarker with a large mean leave-one-out performance loss, low-to-moderate correlation with other biomarkers, and evidence of concentrated contribution to model discrimination;
● complementary biomarker: biomarker with a positive individual contribution and evidence of positive pair excess in at least one relevant biomarker pair;
● redundant biomarker: biomarker with high correlation to another biomarker and negligible or negative mean leave-one-out performance loss;
● weak/stable biomarker: biomarker with relatively stable ranking but limited incremental contribution to performance;
● potentially noisy biomarker: biomarker whose removal was associated with negative mean □ AUC, suggesting that its inclusion may not improve model discrimination in this setting.

These categories were used to summarize the observed signal architecture and were not interpreted as causal or clinically definitive labels.

### 2.12 Uncertainty estimation

Because all structured perturbation analyses were summarized across five outer folds, statistical uncertainty was quantified using bootstrap resampling of fold-level metric values. For each summary metric, 2000 bootstrap resamples with replacement were generated, and the mean value was recalculated in each resample. The 95% confidence interval was then defined as the interval between the 2.5th and 97.5th percentile of the bootstrap distribution.

Bootstrap confidence intervals were computed for:

● leave-one-biomarker-out mean □ AUC,
● pairwise leave-two-out mean □ AUC,
● cumulative ablation performance summaries,
● minimal-panel performance summaries.

For gap-based quantities expressed relative to the full BIOMARKERS model, interval estimates were treated as descriptive approximations. All uncertainty estimates were therefore interpreted conservatively.

### 2.13 Software and reproducibility

All analyses were performed in Python using pandas, NumPy, SciPy, scikit-learn, matplotlib, and seaborn. The study relied on explicit file-based exchange of fold definitions, best hyperparameters, feature lists, and output tables to ensure reproducibility and auditability. Output artifacts included fold-level performance tables, summary tables, correlation maps, redundancy tables, cumulative ablation summaries, minimal-panel summaries, and biomarker typology tables.

The present study used the existing leakage-aware modeling workflow as a fixed analytical scaffold and did not modify the original cohort definition, outer-fold assignments, or winner-model selection procedure.

## 3. Results

### 3.1 Baseline performance of the BIOMARKERS model

The BIOMARKERS variant demonstrated strong discrimination in the baseline outer-fold evaluation, confirming that the biomarker-only feature set contained a substantial predictive signal for STEMI versus NSTEMI classification. Across outer folds, the full biomarker model achieved a mean ROC-AUC of approximately 0.94, with correspondingly high precision-recall performance and low Brier loss (Table 1).

**Table 1.** Baseline performance of the BIOMARKERS model across outer folds.

| outer_fold | roc_auc | pr_auc | brier |
| --- | --- | --- | --- |
| 1 | 0.971 | 0.967 | 0.120 |
| 2 | 0.914 | 0.877 | 0.143 |
| 3 | 0.967 | 0.967 | 0.090 |
| 4 | 1.000 | 1.000 | 0.082 |
| 5 | 0.867 | 0.810 | 0.138 |
| Mean (SD) | 0.944 (0.053) | 0.924 (0.079) | 0.115 (0.028) |

This baseline served as the reference point for all subsequent perturbation analyses. Because the present study was designed to interrogate model structure rather than to re-estimate deployable performance, all downstream analyses focused on changes relative to this fixed biomarker-model baseline.

### 3.2 Correlation structure revealed a highly redundant lipid sub-cluster

Correlation analysis showed marked heterogeneity in the internal structure of the biomarker panel. A strongly collinear lipid cluster was observed, comprising total cholesterol, LDL cholesterol, and non-HDL cholesterol, with pairwise Spearman correlation coefficients reaching approximately 0.93–0.97 (Figure 1, Table S1).

**Figure 1.**
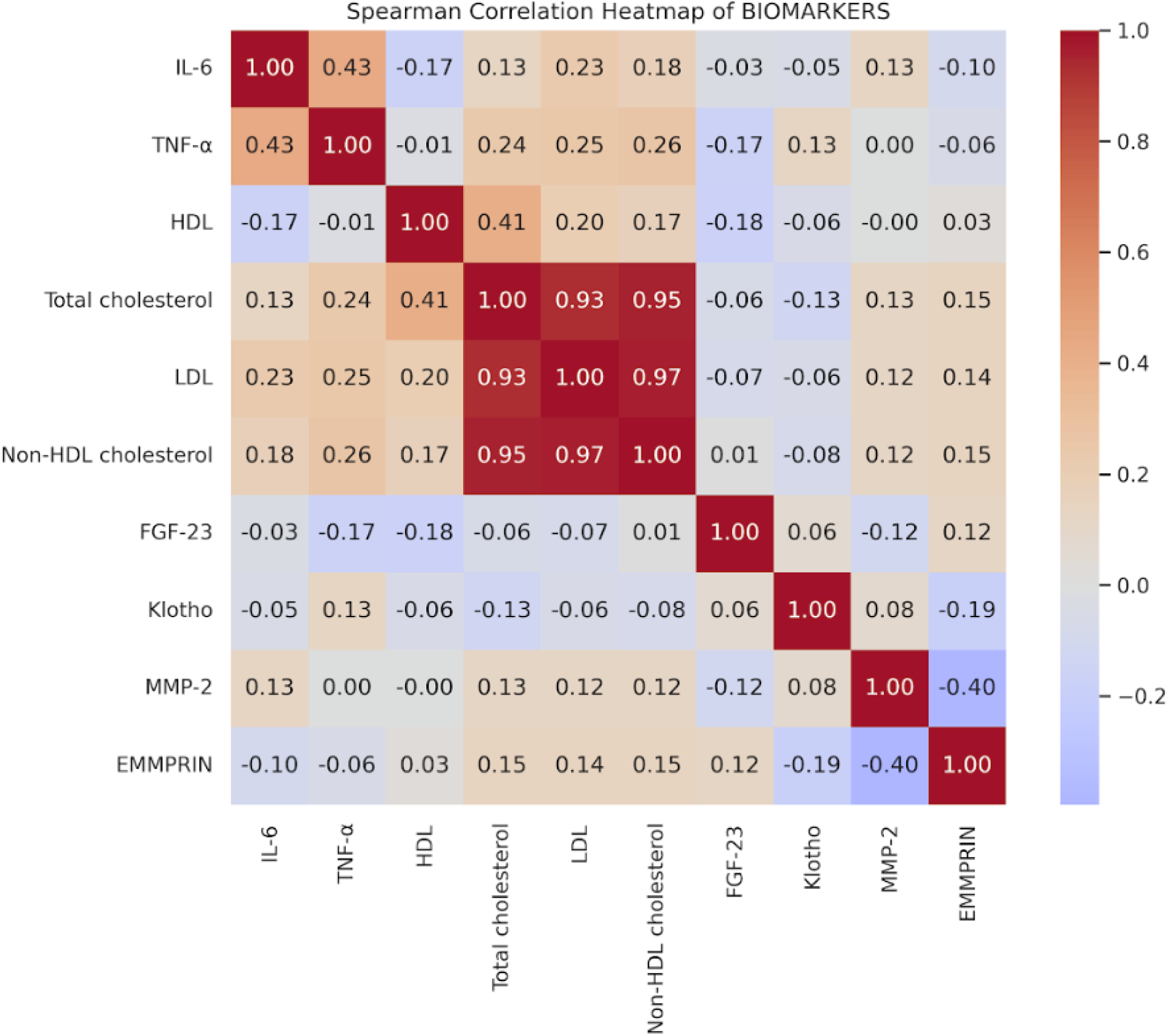
Spearman correlation heatmap of the BIOMARKERS feature set.

Outside this lipid cluster, pairwise correlations were generally lower, suggesting that several non-lipid biomarkers contributed structurally more independent information. In particular, markers related to matrix remodeling and inflammation did not form a similarly tight correlation block.

These findings suggested that the biomarker panel was not organized as a uniformly redundant structure, but rather as a mixture of a highly redundant lipid axis and a more weakly correlated non-lipid component.

### 3.3 Leave-one-biomarker-out analysis identified concentrated dependence on a small subset of biomarkers

Leave-one-biomarker-out analysis demonstrated that model dependence varied markedly across biomarkers (Figure 2, Table 2). Removal of MMP-2 produced the largest average reduction in discrimination, with a mean ΔAUC of approximately 0.16 relative to the full BIOMARKERS model. This effect was substantially larger than for any other single biomarker and was consistently positive across folds, indicating that MMP-2 functioned as a dominant predictive signal within the model structure.

**Figure 2.**
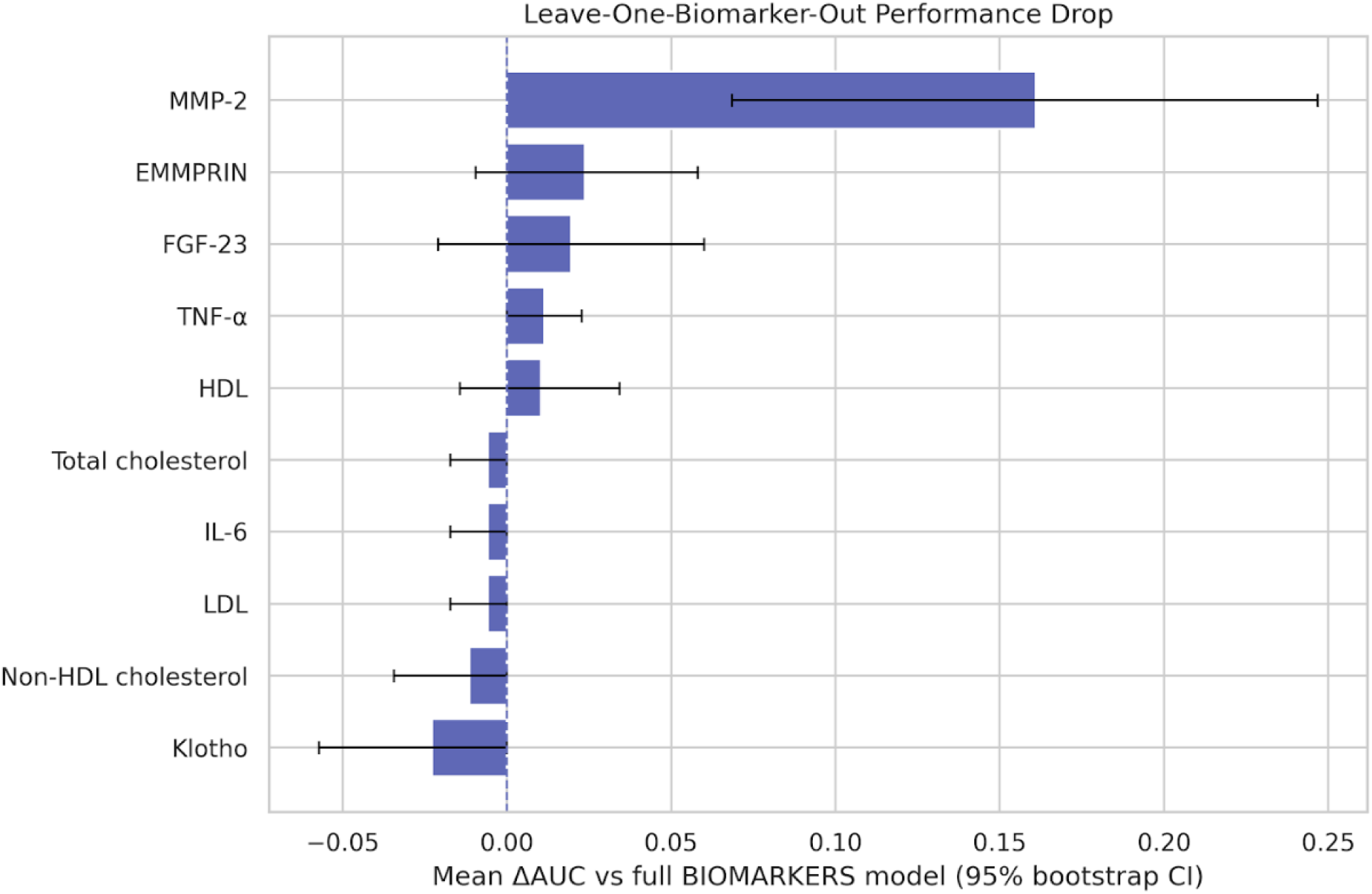
Leave-one-biomarker-out ΔAUC with 95% confidence intervals.

**Table 2.** Leave-one-biomarker-out performance metrics.

| **Biomarker** | **Mean ΔAUC** | **Median ΔAUC** | **SD** | **IQR** | **Folds with harm (%)** |
| --- | --- | --- | --- | --- | --- |
| MMP-2 | 0.161 | 0.171 | 0.109 | 0.067 | 80 |
| EMMPRIN | 0.024 | 0.033 | 0.044 | 0.033 | 60 |
| FGF-23 | 0.02 | 0.029 | 0.052 | 0.083 | 60 |
| TNF-α | 0.011 | 0 | 0.016 | 0.029 | 40 |
| HDL | 0.01 | 0 | 0.034 | 0.029 | 40 |
| Total cholesterol | -0.006 | 0 | 0.013 | 0 | 0 |
| LDL | -0.006 | 0 | 0.013 | 0 | 0 |
| IL-6 | -0.006 | 0 | 0.013 | 0 | 0 |
| Non-HDL cholesterol | -0.011 | 0 | 0.026 | 0 | 0 |
| Klotho | -0.023 | 0 | 0.037 | 0.029 | 0 |

A second tier of biomarkers showed smaller but still positive mean performance losses after removal, including EMMPRIN, FGF-23, and TNF-α. In contrast, several biomarkers showed negligible or inconsistent impact on discrimination when removed.

The lipid-related biomarkers exhibited particularly limited incremental value in the leave-one-out setting, with near-zero or slightly negative ΔAUC values, consistent with redundancy. Removal of Klotho was associated with a slightly negative mean ΔAUC, indicating that average model performance improved marginally when this biomarker was excluded.

Taken together, these results indicated a structurally concentrated signal architecture.

### 3.4 Pairwise leave-two-out analysis distinguished redundancy from conditional complementarity

Pairwise leave-two-out analysis further clarified the internal organization of predictive signals (Figure 3, Figure 4, Table 3).

**Figure 3.**
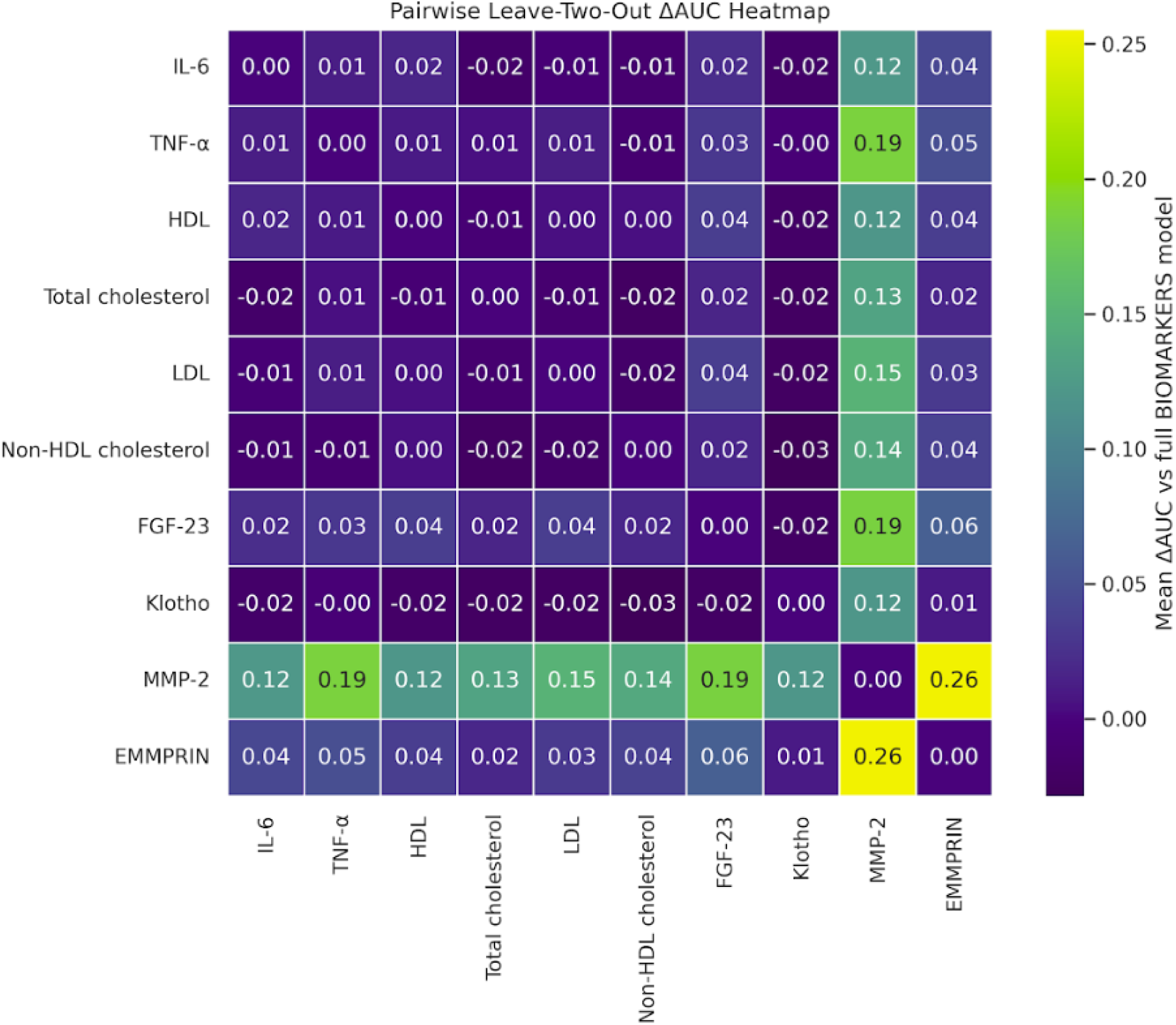
Pairwise leave-two-out ΔAUC heatmap.

**Figure 4.**
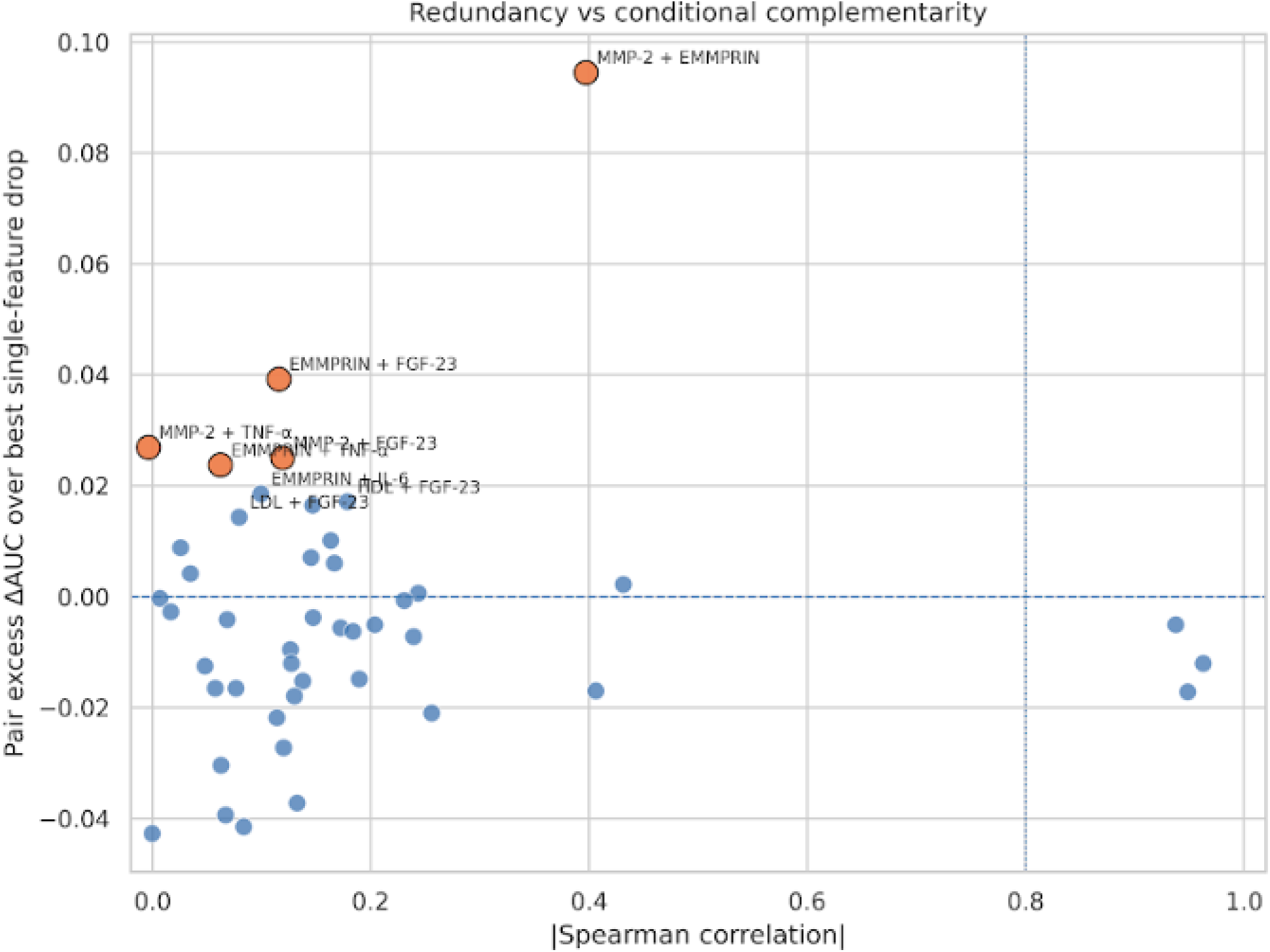
Redundancy vs conditional complementarity plot.

**Table 3.** Pairwise redundancy and conditional complementarity analysis.

| Biomarker 1 | Biomarker 2 | Mean pair ΔAUC | $\vert \rho\vert $ | Pair excess ΔAUC | Interpretation |
| --- | --- | --- | --- | --- | --- |
| MMP-2 | EMMPRIN | 0.255 | 0.4 | 0.094 | potential complementary pair |
| MMP-2 | TNF-α | 0.187 | 0 | 0.026 | potential complementary pair |
| MMP-2 | FGF-23 | 0.186 | 0.12 | 0.025 | potential complementary pair |
| MMP-2 | LDL | 0.15 | 0.12 | -0.01 | partially overlapping pair |
| MMP-2 | Non-HDL cholesterol | 0.139 | 0.12 | -0.022 | partially overlapping pair |
| MMP-2 | Total cholesterol | 0.133 | 0.13 | -0.028 | partially overlapping pair |
| MMP-2 | IL-6 | 0.123 | 0.13 | -0.038 | partially overlapping pair |
| MMP-2 | HDL | 0.12 | 0 | -0.04 | partially overlapping pair |
| MMP-2 | Klotho | 0.12 | 0.08 | -0.041 | partially overlapping pair |
| EMMPRIN | FGF-23 | 0.064 | 0.12 | 0.04 | potential complementary pair |
| EMMPRIN | TNF-α | 0.049 | 0.06 | 0.025 | potential complementary pair |
| EMMPRIN | IL-6 | 0.043 | 0.1 | 0.019 | potential complementary pair |
| EMMPRIN | Non-HDL cholesterol | 0.038 | 0.15 | 0.014 | potential complementary pair |
| LDL | FGF-23 | 0.035 | 0.07 | 0.016 | potential complementary pair |
| HDL | FGF-23 | 0.035 | 0.18 | 0.016 | potential complementary pair |

Simultaneous removal of MMP-2 and EMMPRIN produced the largest mean reduction in model performance (ΔAUC ≈ 0.26), exceeding the stronger of the two individual effects and indicating conditional complementarity.

Several additional biomarker pairs also demonstrated positive pair excess, particularly involving EMMPRIN, FGF-23, and TNF-α. In contrast, highly correlated lipid pairs exhibited minimal additional performance loss beyond single-feature effects, supporting their redundant nature.

### 3.5 Integrated biomarker typology supported a non-uniform signal architecture

Integration of leave-one-out, pairwise, correlation, and stability metrics yielded a coherent biomarker typology (Table 4).

**Table 4.** Integrated biomarker signal typology.

| **Biomarker** | **Stability score** | **Mean ΔAUC** | **Redundancy** | **Signal role** |
| --- | --- | --- | --- | --- |
| MMP-2 | 0.96 | 0.161 | low redundancy | Core driver |
| EMMPRIN | 0.76 | 0.024 | low redundancy | Complementary |
| FGF-23 | 0.49 | 0.02 | low redundancy | Redundant |
| TNF-α | 0.58 | 0.011 | low redundancy | Redundant |
| HDL | 0.13 | 0.01 | low redundancy | Redundant |
| Total cholesterol | 0.3 | -0.006 | low redundancy | Potentially noisy |
| LDL | 0.26 | -0.006 | low redundancy | Potentially noisy |
| IL-6 | 0.38 | -0.006 | low redundancy | Potentially noisy |
| Non-HDL cholesterol | 0.33 | -0.011 | low redundancy | Potentially noisy |
| Klotho | 0.25 | -0.023 | low redundancy | Potentially noisy |

MMP-2 was identified as the primary core driver, while EMMPRIN, FGF-23, and TNF-α functioned as complementary contributors. Lipid biomarkers formed a redundant cluster, and Klotho was consistent with a potentially noisy biomarker.

### 3.6 Cumulative ablation demonstrated progressive model collapse under sequential removal of top-ranked biomarkers

Cumulative ablation analysis showed progressive degradation of model performance as top-ranked biomarkers were removed (Figure 5, Table 5).

**Figure 5.**
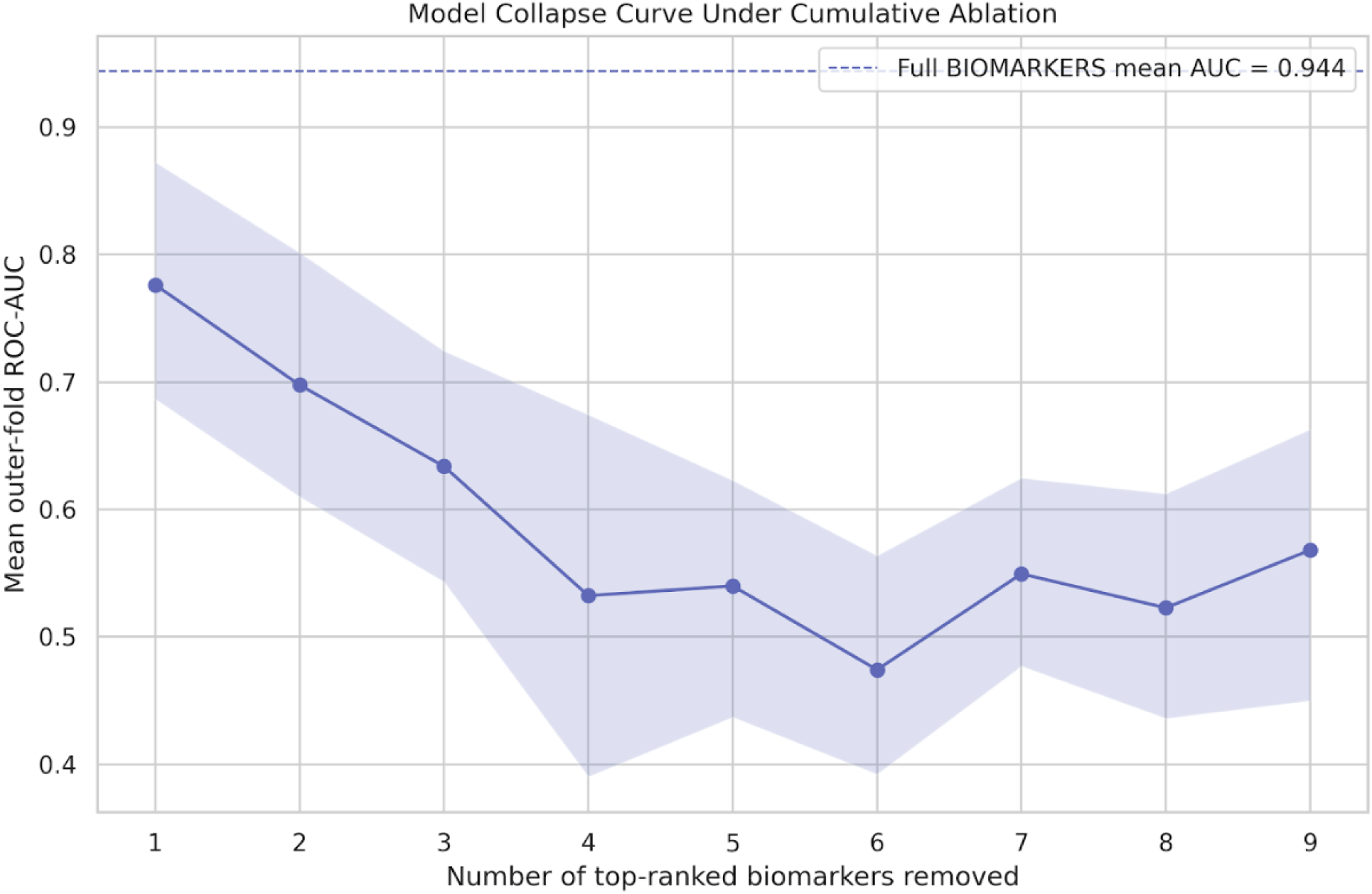
Model collapse curve under cumulative ablation (ROC-AUC with CI)

**Table 5.** Cumulative ablation summary.

| **Number of biomarkers removed** | **Mean ROC-AUC** | **ΔAUC vs full** | **Mean PR-AUC** | **Mean Brier score** |
| --- | --- | --- | --- | --- |
| 1 | 0.776 | 0.168 | 0.782 | 0.185 |
| 2 | 0.698 | 0.246 | 0.685 | 0.235 |
| 3 | 0.634 | 0.31 | 0.622 | 0.256 |
| 4 | 0.532 | 0.411 | 0.603 | 0.284 |
| 5 | 0.54 | 0.404 | 0.56 | 0.287 |
| 6 | 0.474 | 0.47 | 0.506 | 0.309 |
| 7 | 0.55 | 0.394 | 0.59 | 0.278 |
| 8 | 0.523 | 0.421 | 0.59 | 0.293 |
| 9 | 0.568 | 0.376 | 0.634 | 0.295 |

This trajectory indicated strong structural signal concentration.

Removal of the highest-ranked biomarker reduced ROC-AUC from ∼0.94 to ∼0.78, with further decline to near-chance performance after removal of multiple top-ranked features.

### 3.7 Forward reconstruction identified a compact high-yield biomarker panel

Forward panel analysis demonstrated that most predictive signals could be recovered using a small subset of biomarkers (Figure 6, Figure 7, Table 6).

**Figure 6.**
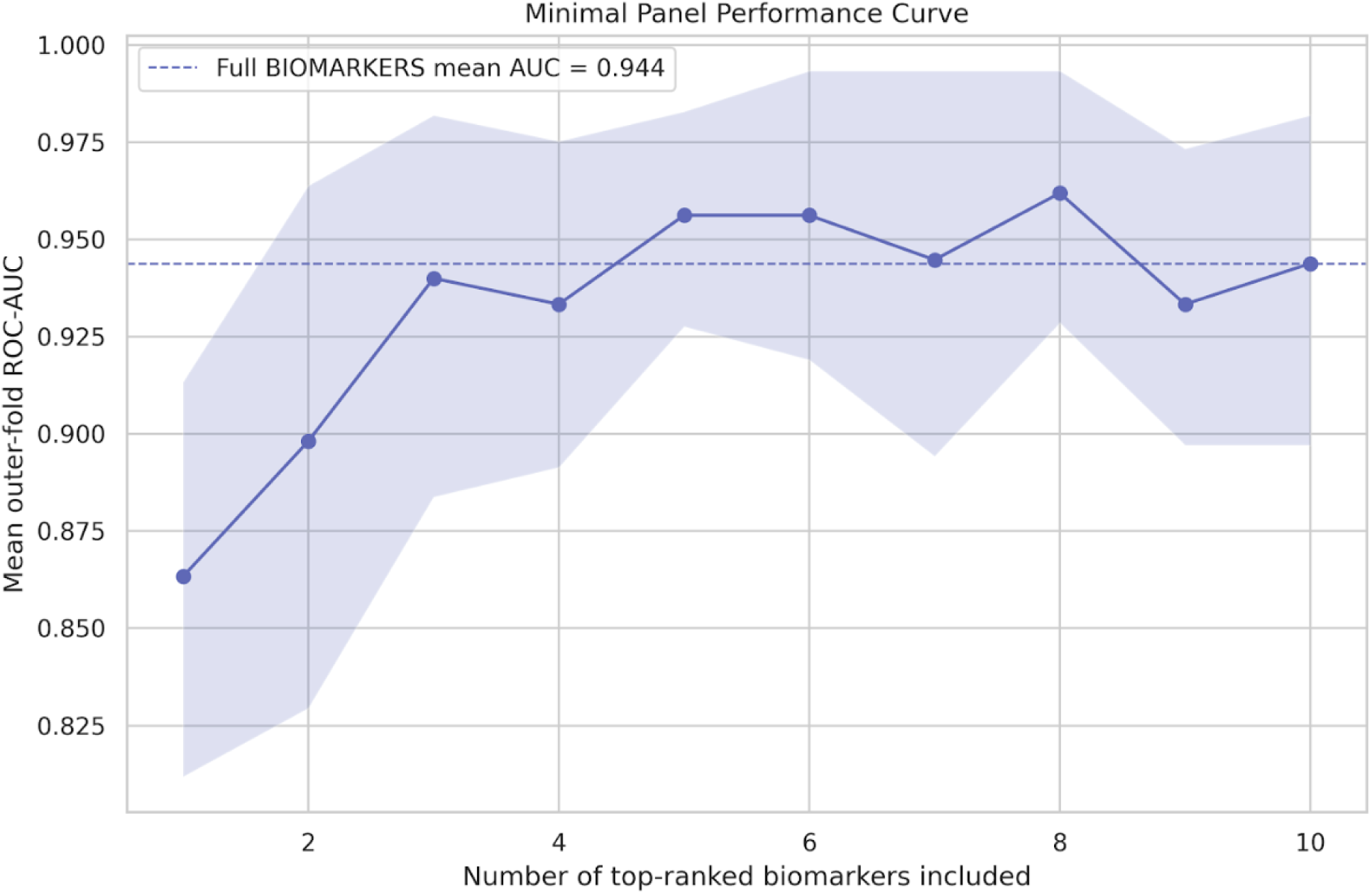
Minimal panel ROC-AUC curve with CI.

**Figure 7.**
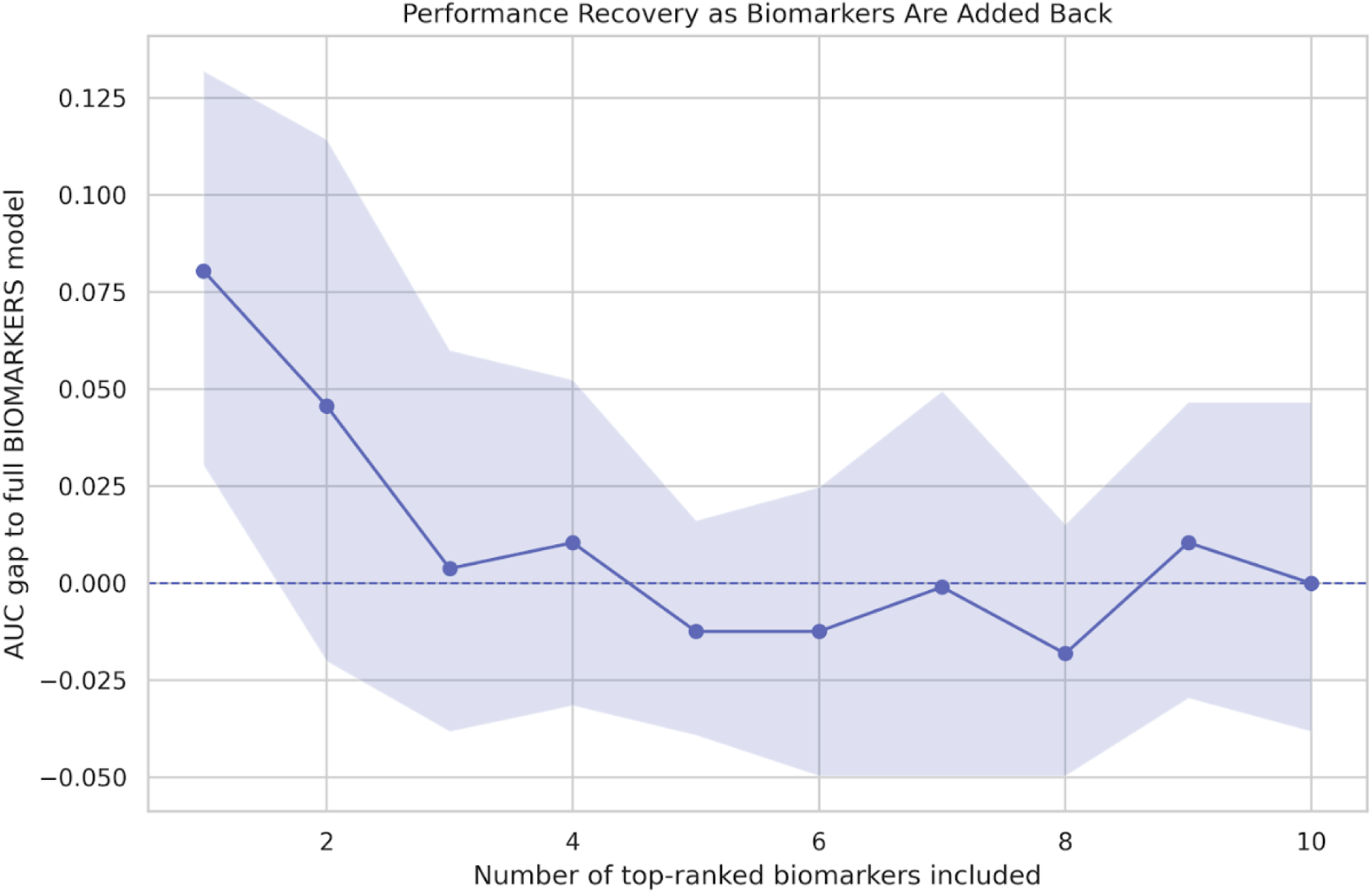
AUC gap to full model vs panel size.

**Table 6.**
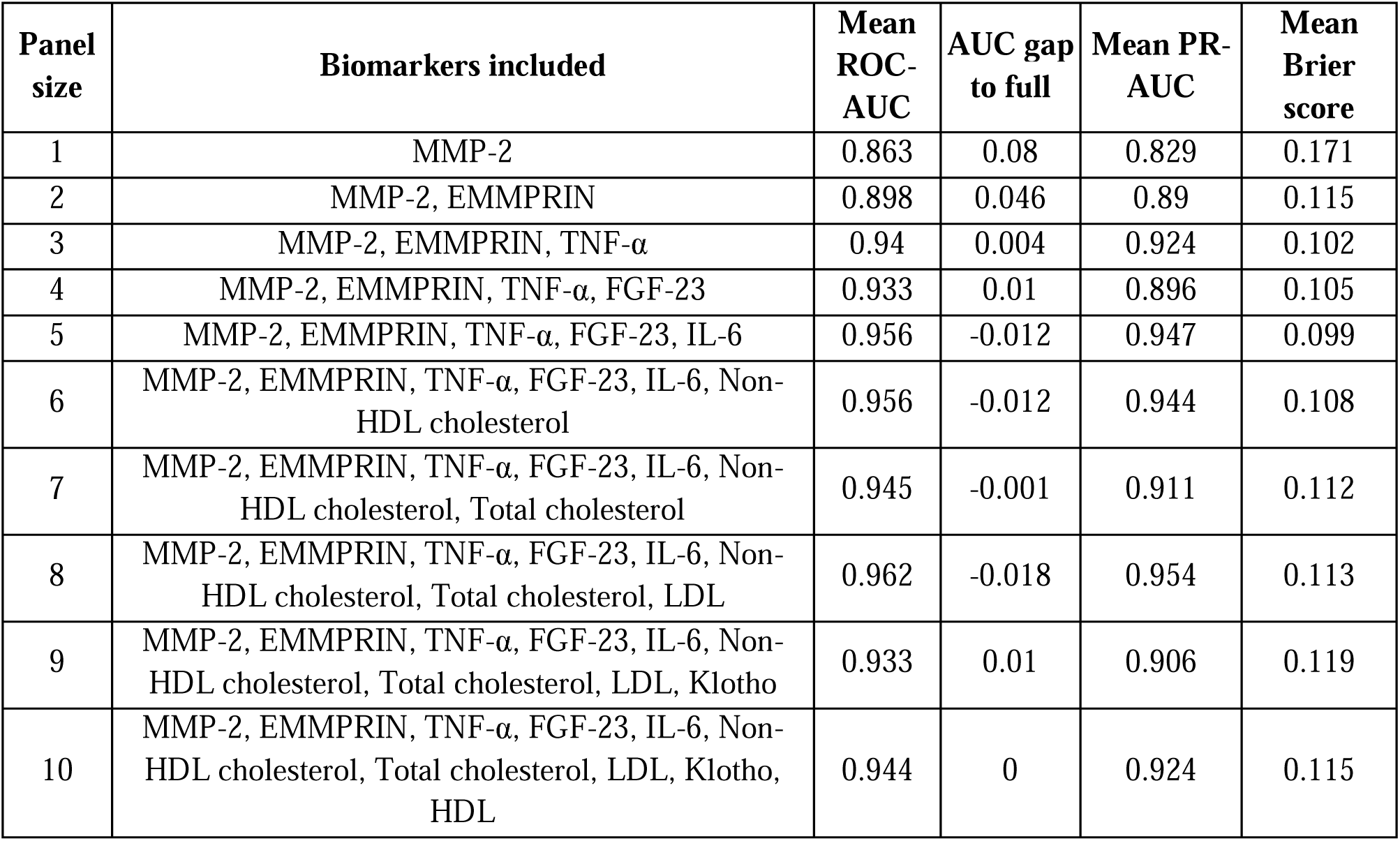
Forward panel performance summary.

| Panel size | Biomarkers included | Mean ROC-AUC | AUC gap to full | Mean PR-AUC | Mean Brier score |
| --- | --- | --- | --- | --- | --- |
| 1 | MMP-2 | 0.863 | 0.08 | 0.829 | 0.171 |
| 2 | MMP-2, EMMPRIN | 0.898 | 0.046 | 0.89 | 0.115 |
| 3 | MMP-2, EMMPRIN, TNF-α | 0.94 | 0.004 | 0.924 | 0.102 |
| 4 | MMP-2, EMMPRIN, TNF-α, FGF-23 | 0.933 | 0.01 | 0.896 | 0.105 |
| 5 | MMP-2, EMMPRIN, TNF-α, FGF-23, IL-6 | 0.956 | -0.012 | 0.947 | 0.099 |
| 6 | MMP-2, EMMPRIN, TNF-α, FGF-23, IL-6, Non-HDL cholesterol | 0.956 | -0.012 | 0.944 | 0.108 |
| 7 | MMP-2, EMMPRIN, TNF-α, FGF-23, IL-6, Non-HDL cholesterol, Total cholesterol | 0.945 | -0.001 | 0.911 | 0.112 |
| 8 | MMP-2, EMMPRIN, TNF-α, FGF-23, IL-6, Non-HDL cholesterol, Total cholesterol, LDL | 0.962 | -0.018 | 0.954 | 0.113 |
| 9 | MMP-2, EMMPRIN, TNF-α, FGF-23, IL-6, Non-HDL cholesterol, Total cholesterol, LDL, Klotho | 0.933 | 0.01 | 0.906 | 0.119 |
| 10 | MMP-2, EMMPRIN, TNF-α, FGF-23, IL-6, Non-HDL cholesterol, Total cholesterol, LDL, Klotho, HDL | 0.944 | 0 | 0.924 | 0.115 |

A three-biomarker panel (MMP-2, EMMPRIN, TNF-α) achieved performance nearly identical to the full model (ΔAUC ≈ 0.004).

Beyond this point, performance gains were modest and non-monotonic, consistent with a diminishing-returns structure.

### 3.8 Summary of signal architecture findings

Taken together, the analyses supported a structured biomarker signal architecture characterized by:

● a dominant core driver (MMP-2),
● a small set of complementary biomarkers,
● a redundant lipid cluster,
● and a residual group of weak or noisy features.

This pattern indicates that model performance arises from a compact and unevenly distributed signal structure rather than uniform contribution across biomarkers.

## 4. Discussion

### 4.1 Principal findings

In this study, we performed a structured analysis of the biomarker signal architecture within a machine-learning model for STEMI versus NSTEMI classification. Rather than focusing on overall model performance, we examined how predictive signal was distributed across biomarkers and how this structure influenced model behavior under systematic perturbation.

Three principal findings emerged. First, the predictive signal was highly concentrated, with a single dominant biomarker (MMP-2) and a small group of additional contributors accounting for most of the model’s discrimination capacity. Second, the biomarker panel exhibited a clear internal structure composed of a redundant lipid sub-cluster and a conditionally complementary non-lipid substructure. Third, most predictive performance could be retained using a compact biomarker subset, using a compact biomarker subset, consistent with the existence of a minimal high-yield panel, although no single panel configuration could be considered uniquely optimal.

Taken together, these results suggest that biomarker importance in this setting is best understood not as an isolated ranking problem, but as an emergent property of interacting signal components within a structured system.

### 4.2 Structural signal concentration and the dominance of MMP-2

The leave-one-biomarker-out analysis demonstrated a marked asymmetry in feature importance, with MMP-2 exerting a substantially larger impact on model performance than any other biomarker. This observation is consistent with a core-driver architecture, in which a single variable captures a large proportion of the discriminative signal.

From a methodological perspective, such concentration has important implications. Models that rely disproportionately on a single feature may achieve high apparent performance but can be structurally fragile, particularly if that feature is noisy, poorly standardized, or unavailable in external datasets. At the same time, the presence of a dominant predictive signal does not necessarily imply redundancy of the remaining features, as demonstrated by the complementary roles observed in pairwise analyses.

From a biological standpoint, MMP-2 is linked to extracellular matrix remodeling and post-infarction tissue dynamics, which are mechanistically relevant to myocardial injury. However, the present study was not designed to establish causal importance. Accordingly, the observed dominance of MMP-2 should be interpreted as a property of model-dependent signal capture, not as definitive evidence of clinical primacy.

### 4.3 Redundancy versus conditional complementarity

A key contribution of this work is the explicit separation of redundancy from conditional complementarity, two concepts that are often conflated in feature-importance analyses.

The lipid-related biomarkers (total cholesterol, LDL, non-HDL cholesterol) formed a highly correlated cluster and demonstrated minimal incremental contribution when evaluated either individually or in pairs. This pattern is consistent with redundant representation of overlapping physiological information, where multiple variables encode similar aspects of lipid metabolism.

In contrast, several non-lipid biomarkers, particularly EMMPRIN, FGF-23, and TNF-α, exhibited conditional complementarity, as evidenced by positive pair excess beyond single-feature effects. This indicates that these biomarkers may contribute information that becomes relevant only in the presence of other variables, reflecting interaction-like behavior within the model level.

These findings highlight an important limitation of conventional univariable or additive importance metrics: they may underestimate the role of features whose contribution depends on the broader feature context. The distinction between redundancy and complementarity therefore provides a more nuanced framework for interpreting biomarker relevance in multivariable machine-learning systems.

### 4.4 Minimal high-yield biomarker panels and diminishing returns

The forward panel analysis demonstrated that most of the model’s predictive performance could be retained using a small subset of top-ranked biomarkers. In particular, a three-biomarker panel achieved performance nearly indistinguishable from the full model, suggesting that the majority of the signal was concentrated in a compact subset.

Importantly, performance gains beyond this subset were modest and non-monotonic, consistent with a diminishing-returns structure. This pattern argues against the assumption that adding more biomarkers will necessarily improve predictive performance in a stable or clinically meaningful way.

However, these findings should not be interpreted as identifying a clinically optimal or deployable biomarker panel. The forward selection procedure was anchored to a predefined ranking and was intended as a diagnostic exploration of signal recovery, not as an optimization strategy. External validation, clinical feasibility, and assay standardization would all be required before proposing any reduced panel for practical use.

### 4.5 Implications for biomarker-based machine learning in cardiology

The results of this study have broader implications for the design and interpretation of biomarker-based machine-learning models in cardiovascular medicine.

First, they suggest that model performance alone is insufficient to characterize a biomarker system. Two models with similar AUC values may rely on fundamentally different signal architectures, with implications for robustness, interpretability, and generalizability.

Second, they support the concept that biomarker panels should be evaluated not only in terms of aggregate performance but also in terms of structural properties, including redundancy, complementarity, and signal concentration. Such properties may influence how models behave under dataset shift, missing data, or partial feature availability.

Third, the identification of redundant clusters highlights opportunities for feature simplification, whereas the detection of complementary relationships suggests that certain combinations of biomarkers may provide synergistic information that is not apparent in isolation.

Finally, the framework used in this study—combining leave-analysis, pairwise perturbation, and cumulative ablation—may apply to other clinical domains where understanding model structure is as important as optimizing performance.

### 4.6 Limitations

Several limitations should be considered when interpreting these findings.

First, the analysis was conducted on a relatively small dataset (n = 152), which limits statistical precision and increases the risk of overinterpretation of small performance differences. Although bootstrap-derived confidence intervals were used to quantify uncertainty, these estimates should be interpreted cautiously.

Second, all analyses were based on internal cross-validation without external validation. As a result, the observed signal architecture may be specific to this dataset and may not generalize to other populations or clinical settings.

Third, the ranking used for cumulative ablation and forward panel analyses was derived from a predefined stability metric and was not re-optimized within the present study. While this approach reduces the risk of circular analysis, it also means that the resulting panels should be interpreted as diagnostic constructs rather than optimized solutions.

Fourth, the operational classification of biomarkers into roles such as “core driver” or “complementary contributor” was based on model-derived metrics rather than biological validation. These categories are therefore descriptive and should not be interpreted as mechanistic or causal classifications.

Finally, the modeling framework relied on specific algorithms and preprocessing choices defined in the original workflow. Although these choices were held constant to preserve methodological consistency, different modeling strategies might yield different signal architectures.

### 4.7 Conclusions

In this machine-learning analysis of post–myocardial infarction biomarkers, predictive signals were found to be unevenly distributed and structurally organized, rather than uniformly shared across features. The model was characterized by a dominant core biomarker, a small set of conditionally complementary contributors, and a redundant lipid cluster with limited incremental value.

These findings support the concept of biomarker signal architecture as a useful framework for understanding how predictive models operate beyond aggregate performance metrics. In particular, they suggest that model behavior is shaped by the interaction of concentrated signal components, rather than by independent contributions of individual biomarkers.

Future studies should investigate whether such structural patterns are reproducible across larger cohorts and whether they can inform the development of more robust, interpretable, and clinically actionable machine-learning models in cardiovascular medicine.

## Data Availability

The analytical code used in this study is publicly available in a dedicated GitHub repository: https://github.com/npiorkowska-science/biomarker-signal-architecture-mi-ml. The clinical dataset contains sensitive patient data and is not publicly available. Aggregated results and derived data supporting the findings of this study are available from the corresponding author upon reasonable request.

## Data and Code Availability

The analytical code used in this study is publicly available in a dedicated GitHub repository: https://github.com/npiorkowska-science/biomarker-signal-architecture-mi-ml

## Funding

This research received no specific grant from any funding agency in the public, commercial, or not-for-profit sectors.

## Conflict of Interest

The authors declare that they have no conflicts of interest relevant to this study.

## Ethics Statement

The study was conducted in accordance with the Declaration of Helsinki. The use of clinical data for the proof-of-concept analysis was approved by the Ethics Committee of the Medical University of Wroclaw (approval numbers: KB-54/2019, KB-514/2019, KB-387/2021). All data used in the analysis were anonymized prior to processing. The systematic review part of the study was conducted using previously published studies and did not require additional ethical approval.

## Author Contributions

Natalia Piórkowska conceptualized the study, designed the methodology, performed the analysis, interpreted the results, and wrote the manuscript.

Agnieszka Olejnik: Methodology (clinical dataset design and biomarker panel selection), Data curation, Investigation, Validation, Writing – original draft, Writing – review & editing.

Alan Ostromęcki conceptualized the study, designed the methodology, visualization and wrote the manuscript.

Wiktor Kuliczkowski: The acquisition and curation of patient clinical and biomarker data.

Andrzej Mysiak: The acquisition and curation of patient clinical and biomarker data.

Iwona Bil-Lula: Methodology (clinical dataset design and biomarker panel selection), Data curation, Investigation, Validation, Writing – review & editing.

## Acknowledgments

The authors acknowledge the Wroclaw Medical University for providing access to the clinical data used in the proof-of-concept analysis. The authors also thank collaborators who contributed to the development of the methodological framework and provided valuable feedback during the preparation of this study.

## Supplementary materials

**Table S1**. Pairwise Spearman correlation coefficients and redundancy classification for all biomarker pairs.

## Notes

### Competing Interest Statement

The authors have declared no competing interest.

## References

[1] R.A. Byrne, X. Rossello, J.J. Coughlan, E. Barbato, C. Berry, A. Chieffo, M.J. Claeys, G.-A. Dan, M.R. Dweck, M. Galbraith, M. Gilard, L. Hinterbuchner, E.A. Jankowska, P. Jüni, T. Kimura, V. Kunadian, M. Leosdottir, R. Lorusso, R.F.E. Pedretti, A.G. Rigopoulos, M. Rubini Gimenez, H. Thiele, P. Vranckx, S. Wassmann, N.K. Wenger, B. Ibanez, ESC Scientific Document Group, 2023 ESC Guidelines for the management of acute coronary syndromes, Eur. Heart J. 44 (2023) 3720–3826. 10.1093/eurheartj/ehad191.

[2] C.C.F.M.J. Baaten, M. Nagy, W. Bergmeier, H.M.H. Spronk, P.E.J. van der Meijden, Platelet biology and function: plaque erosion vs. rupture, Eur. Heart J. 45 (2024) 18–31. 10.1093/eurheartj/ehad720.

[3] M.A. Matter, F. Paneni, P. Libby, S. Frantz, B.E. Stähli, C. Templin, A. Mengozzi, Y.-J. Wang, T.M. Kündig, L. Räber, F. Ruschitzka, C.M. Matter, Inflammation in acute myocardial infarction: the good, the bad and the ugly, Eur. Heart J. 45 (2024) 89–103. 10.1093/eurheartj/ehad486.

[4] J. Budzianowski, D. Hiczkiewicz, H. Ficner, J. Rzeźniczak, M. Słomczyński, D. Kasprzak, J. Hiczkiewicz, P. Burchardt, Updated focused review of hematological, inflammatory, and lipid biomarkers in acute coronary syndrome, Arch. Med. Sci. 21 (2025) 2258–2266. 10.5114/aoms/208106.

[5] A. Aleksova, A.L. Fluca, A.P. Beltrami, E. Dozio, G. Sinagra, M. Marketou, M. Janjusevic, Biomarkers of Importance in Monitoring Heart Condition After Acute Myocardial Infarction, J. Clin. Med. 14 (2025) 129. 10.3390/jcm14010129.

[6] B. Toprak, J. Weimann, J. Lehmacher, P.M. Haller, T.S. Hartikainen, A. Schock, M. Karakas, T. Renné, T. Zeller, R. Twerenbold, N.A. Sörensen, D. Westermann, J.T. Neumann, Prognostic utility of a multi-biomarker panel in patients with suspected myocardial infarction, Clin. Res. Cardiol. Off. J. Ger. Card. Soc. 113 (2024) 1682–1691. 10.1007/s00392-023-02345-7.

[7] C. Wies, R. Miltenberger, G. Grieser, A. Jahn-Eimermacher, Exploring the variable importance in random forests under correlations: a general concept applied to donor organ quality in post-transplant survival, BMC Med. Res. Methodol. 23 (2023) 209. 10.1186/s12874-023-02023-2.

[8] T. Liu, A. Krentz, L. Lu, V. Curcin, Machine learning based prediction models for cardiovascular disease risk using electronic health records data: systematic review and meta-analysis, Eur. Heart J. Digit. Health 6 (2025) 7–22. 10.1093/ehjdh/ztae080.

[9] H. Climente-González, M. Oh, U. Chajewska, R. Hosseini, S. Mukherjee, W. Gan, M. Traylor, S. Hu, G. Fatemifar, J. Ghouse, P.P. Del Villar, E. Vernet, N. Koelling, L. Du, R. Abraham, C. Li, J.M.M. Howson, Interpretable machine learning leverages proteomics to improve cardiovascular disease risk prediction and biomarker identification, Commun. Med. 5 (2025) 170. 10.1038/s43856-025-00872-0.

[10] A. Niazai, H. Jamil, M. Hameed, S. Sheikh, M.R. Nisar, Artificial intelligence in cardiovascular diagnostics: a systematic review and descriptive analysis of clinical applications and diagnostic performance, BMC Cardiovasc. Disord. 25 (2025) 849. 10.1186/s12872-025-05327-x.

[11] M. Fu, R. He, Z. Zhang, F. Ma, L. Shen, Y. Zhang, M. Duan, Y. Zhang, Y. Wang, L. Zhu, J. He, Multinomial machine learning identifies independent biomarkers by integrated metabolic analysis of acute coronary syndrome, Sci. Rep. 13 (2023) 20535. 10.1038/s41598-023-47783-5.

[12] A.M. Salih, I.B. Galazzo, P. Gkontra, E. Rauseo, A.M. Lee, K. Lekadir, P. Radeva, S.E. Petersen, G. Menegaz, A review of evaluation approaches for explainable AI with applications in cardiology, Artif. Intell. Rev. 57 (2024) 240. 10.1007/s10462-024-10852-w.

[13] Y. Yang, C.-Y. Liao, E. Keyvanshokooh, H. Shao, M.B. Weber, F.J. Pasquel, G.-G.P. Garcia, A Responsible Framework for Assessing, Selecting, and Explaining Machine Learning Models in Cardiovascular Disease Outcomes Among People With Type 2 Diabetes: Methodology and Validation Study, JMIR Med. Inform. 13 (2025) e66200. 10.2196/66200.

[14] M.L. Lindsey, A.F. Oliver, A. Gaye, P.N. Nde, K.Y. DeLeon-Pennell, G.E. González, Cellular Interactions of Cardiac Repair After Myocardial Infarction, Cells 14 (2025) 1903. 10.3390/cells14231903.

[15] A. Imbesi, A. Greco, M. Spagnolo, C. Laudani, C. Raffo, S. Finocchiaro, P.M. Mazzone, D. Landolina, M.S. Mauro, L. Cutore, G. Di Leo, D.C. Faro, N. Ammirabile, D. Giacoppo, D. Capodanno, Targeting Inflammation After Acute Myocardial Infarction, JACC 86 (2025) 1146–1169. 10.1016/j.jacc.2025.07.064.

[16] H. Bräuninger, S. Krüger, L. Bacmeister, A. Nyström, K. Eyerich, D. Westermann, D. Lindner, Matrix metalloproteinases in coronary artery disease and myocardial infarction, Basic Res. Cardiol. 118 (2023) 18. 10.1007/s00395-023-00987-2.

[17] M. Wolosowicz, S. Prokopiuk, T.W. Kaminski, The Complex Role of Matrix Metalloproteinase-2 (MMP-2) in Health and Disease, Int. J. Mol. Sci. 25 (2024) 13691. 10.3390/ijms252413691.

[18] A. Krzywonos-Zawadzka, A. Franczak, A. Olejnik, M. Radomski, J.F. Gilmer, G. Sawicki, M. Woźniak, I. Bil-Lula, Cardioprotective effect of MMP-2-inhibitor-NO-donor hybrid against ischaemia/reperfusion injury, J. Cell. Mol. Med. 23 (2019) 2836–2848. 10.1111/jcmm.14191.

[19] I. Cuadrado, M.J.G.M. Piedras, I. Herruzo, M. del C. Turpin, B. Castejón, P. Reventun, A. Martin, M. Saura, J.L. Zamorano, C. Zaragoza, EMMPRIN-Targeted Magnetic Nanoparticles for In Vivo Visualization and Regression of Acute Myocardial Infarction, Theranostics 6 (2016) 545–557. 10.7150/thno.13352.

[20] N.N. Mehta, E. deGoma, M.D. Shapiro, IL-6 and Cardiovascular Risk: A Narrative Review, Curr. Atheroscler. Rep. 27 (2024) 12. 10.1007/s11883-024-01259-7.

[21] H. Li, Y. Bian, Fibroblast-derived interleukin-6 exacerbates adverse cardiac remodeling after myocardial infarction, Korean J. Physiol. Pharmacol. Off. J. Korean Physiol. Soc. Korean Soc. Pharmacol. 28 (2024) 285–294. 10.4196/kjpp.2024.28.3.285.

[22] D.M. Brie, C. Morno, O. Adam, A. Tîrziu, R. Popescu, A.D. Brie, Inflammatory Mechanisms in Acute Coronary Syndromes: From Pathophysiology to Therapeutic Targets, Cells 15 (2026) 72. 10.3390/cells15010072.

[23] P.M. Ridker, N. Rifai, M. Pfeffer, F. Sacks, S. Lepage, E. Braunwald, Elevation of Tumor Necrosis Factor-α and Increased Risk of Recurrent Coronary Events After Myocardial Infarction, Circulation 101 (2000) 2149–2153. 10.1161/01.CIR.101.18.2149.

[24] L. Kong, Z.-B. Yang, X.-H. Chen, X.-Q. Quan, H.-T. Liu, A.-P. Qiu, The causal relationship between triglycerides and myocardial infarction: A two-sample Mendelian randomization, Medicine (Baltimore) 103 (2024) e39595. 10.1097/MD.0000000000039595.

[25] A. Nowowiejska-Wiewióra, K. Wita, Z. Mędrala, L. Tomkiewicz-Pająk, K. Bujak, K. Mizia-Stec, P. Brzychczy, M. Gąsior, Z. Gąsior, A. Kulbat, Z. Kalarus, W. Wojakowski, P. Trzeciak, A. Witkowski, M. Banach, J. Legutko, Dyslipidemia treatment and attainment of LDL-cholesterol treatment goals in patients participating in the Managed Care for Acute Myocardial Infarction Survivors program, Pol. Heart J. Kardiologia Pol. 81 (2023) 359–365. 10.33963/KP.a2023.0045.

[26] A. Zeliaś, A. Bednarek, D. Dykla, R. Wysocka, P. Szczygiel, D. Dudek, Lipid goal achievement one month after myocardial infarction: Observational real-world study from the Polish population, Pol. Heart J. Kardiologia Pol. 82 (2024) 892–894. 10.33963/v.phj.101554.

[27] D. Sehayek, J. Cole, E. Björnson, J.T. Wilkins, M.B. Mortensen, L. Dufresne, K.M. Pencina, M.J. Pencina, G. Thanassoulis, A.D. Sniderman, ApoB, LDL-C, and non-HDL-C as markers of cardiovascular risk, J. Clin. Lipidol. 19 (2025) 844–859. 10.1016/j.jacl.2025.05.024.

[28] J. Płonka, A. Olejnik, A. Klus, E. Gawrylak-Dryja, N. Wężyk, L. Rzepiela, K. Dąbrowska, I. Bil-Lula, M. Gierlotka, sαKlotho protein is a useuful biomarker and a promising cardioprotective agent in acute heart failure, Arch. Med. Sci. (2024). 10.5114/aoms/192239.

[29] J. Płonka, A. Olejnik, A. Klus, E. Gawrylak-Dryja, N. Wężyk, L. Rzepiela, K. Dąbrowska, K. Nalewajko, T. Porażko, I. Bil-Lula, M. Gierlotka, Use of FGF-23 and sαKlotho for Risk Stratification in Patients with Acute Heart Failure, J. Clin. Med. 14 (2025) 860. 10.3390/jcm14030860.

[30] A. Olejnik, J. Płonka, W. Kuliczkowski, A. Mysiak, M. Gierlotka, I.D. Bil-Lula, Diagnostic potential of increased Klotho and FGF23 protein concentrations after myocardial infarction in patients with acute coronary syndrome, Cardiol. J. 0 (2025). 10.5603/cj.98861.

[31] A.S. Mohd Faizal, W.Y. Hon, T.M. Thevarajah, S.M. Khor, S.-W. Chang, A biomarker discovery of acute myocardial infarction using feature selection and machine learning, Med. Biol. Eng. Comput. (2023) 1–15. 10.1007/s11517-023-02841-y.

[32] J.H. Jeong, K.-S. Lee, S.-M. Park, S.R. Kim, M.-N. Kim, S.C. Chae, S.-H. Hur, I.W. Seong, S.K. Oh, T.H. Ahn, M.H. Jeong, Prediction of longitudinal clinical outcomes after acute myocardial infarction using a dynamic machine learning algorithm, Front. Cardiovasc. Med. 11 (2024). 10.3389/fcvm.2024.1340022.

[33] Y. Fan, M. Liu, G. Sun, An interpretable machine learning framework for diagnosis and prognosis of COVID-19, PLOS ONE 18 (2023) e0291961. 10.1371/journal.pone.0291961.

[34] D.P. Upadhyaya, Y. Tarabichi, K. Prantzalos, S. Ayub, D.C. Kaelber, S.S. Sahoo, Machine Learning Interpretability Methods to Characterize the Importance of Hematologic Biomarkers in Prognosticating Patients with Suspected Infection, Comput. Biol. Med. 183 (2024) 109251. 10.1016/j.compbiomed.2024.109251.

[35] N. Piórkowska, A. Olejnik, L. Madeyski, A. Musz, W. Kuliczkowski, A. Mysiak, A. Żyłka, I. Bil-Lula, Artificial Intelligence for Cardiac Biomarkers After Myocardial Infarction: A Systematic Review and a Leakage-Aware Modeling Framework, medRxiv (2026) 2026.04.28.26351958. 10.64898/2026.04.28.26351958.

